# Benchmark of thirteen bioinformatic pipelines for metagenomic virus diagnostics using datasets from clinical samples

**DOI:** 10.1101/2021.05.04.21256618

**Authors:** Jutte J.C. de Vries, Julianne R. Brown, Nicole Fischer, Igor A. Sidorov, Sofia Morfopoulou, Jiabin Huang, Bas B. Oude Munnink, Arzu Sayiner, Alihan Bulgurcu, Christophe Rodriguez, Guillaume Gricourt, Els Keyaerts, Leen Beller, Claudia Bachofen, Jakub Kubacki, Samuel Cordey, Florian Laubscher, Dennis Schmitz, Martin Beer, Dirk Hoeper, Michael Huber, Verena Kufner, Maryam Zaheri, Aitana Lebrand, Anna Papa, Sander van Boheemen, Aloys C.M. Kroes, Judith Breuer, F. Xavier Lopez-Labrador, Eric C.J. Claas, on behalf of the ESCV Network on Next-Generation Sequencing

## Abstract

Metagenomic sequencing is increasingly being used in clinical settings for difficult to diagnose cases. The performance of viral metagenomic protocols relies to a large extent on the bioinformatic analysis. In this study, the European Society for Clinical Virology (ESCV) Network on NGS (ENNGS) initiated a benchmark of metagenomic pipelines currently used in clinical virological laboratories.

**Methods:** Metagenomic datasets from 13 clinical samples from patients with encephalitis or viral respiratory infections characterized by PCR were selected. The datasets were analysed with 13 different pipelines currently used in virological diagnostic laboratories of participating ENNGS members. The pipelines and classification tools were: Centrifuge, DAMIAN, DIAMOND, DNASTAR, FEVIR, Genome Detective, Jovian, MetaMIC, MetaMix, One Codex, RIEMS, VirMet, and Taxonomer. Performance, characteristics, clinical use, and user-friendliness of these pipelines were analysed.

**Results:** Overall, viral pathogens with high loads were detected by all the evaluated metagenomic pipelines. In contrast, lower abundance pathogens and mixed infections were only detected by 3/13 pipelines, namely DNASTAR, FEVIR, and MetaMix. Overall sensitivity ranged from 80% (10/13) to 100% (13/13 datasets). Overall positive predictive value ranged from 71-100%. The majority of the pipelines classified sequences based on nucleotide similarity (8/13), only a minority used amino acid similarity, and 6 of the 13 pipelines assembled sequences *de novo*. No clear differences in performance were detected that correlated with these classification approaches. Read counts of target viruses varied between the pipelines over a range of 2-3 log, indicating differences in limit of detection.

**Conclusion:** A wide variety of viral metagenomic pipelines is currently used in the participating clinical diagnostic laboratories. Detection of low abundant viral pathogens and mixed infections remains a challenge, implicating the need for standardization and validation of metagenomic analysis for clinical diagnostic use. Future studies should address the selective effects due to the choice of different reference viral databases.

## Introduction

Viral metagenomic next-generation sequencing (mNGS) is increasingly being used in virology laboratories for the diagnosis of patients with suspected but unexplained infectious diseases. The current main clinical application of viral metagenomics is for diagnosing encephalitis of unknown cause [1, 2], but metagenomic sequencing is considered useful in a growing number of other clinical syndromes [3-6]. Although many wet-lab challenges need to be faced as well [14], the performance of metagenomic methods is largely dependent on accurate bioinformatic analysis, and both classification algorithms and databases are crucial factors determining the overall performance of the pipelines [7] [55]. A wide range of metagenomic pipelines and taxonomic classifiers have been developed, commonly for the purpose of biodiversity studies analysing the composition of the microbiome in different cohorts. In contrast, when applying metagenomics to patient diagnostics, potential false-negative and false-positive bioinformatic classification results can have significant consequences for patient care. Most reports on bioinformatic tools for metagenomic analysis for virus diagnostics typically describe algorithms and validations of single in-house developed pipelines developed by the authors themselves [8-12]. Most reports on bioinformatic tools for metagenomic analysis for virus diagnostics typically describe algorithms and validations of single in-house developed pipelines developed by the authors themselves [13], and recently a metagenomic benchmarking trial among Swiss virology laboratories has been conducted [7]. Recently, ESCV Network on NGS (ENNGS) recommendations for the introduction of next-generation sequencing in clinical virology, part II: bioinformatic analysis and reporting were published [55], aiming to address the challenges involved. While a professional External Quality Assessment (EQA) program is currently in preparation by Quality Control for Molecular Diagnostics (QCMD), the ENNGS [14] [55] conducted the presented benchmark of bioinformatic pipelines of the participating diagnostic laboratories using viral metagenomic datasets derived from clinical samples, in order to assist laboratories with selection and optimization of tools to be implemented for clinical use.

## Methods

### Datasets

To exclude differences in wet-lab procedures, the same raw, untrimmed metagenomic datasets were provided, so that the participants had standardized datasets for bioinformatic analysis.

In total, 13 clinical metagenomic datasets from samples well-characterized by RT-PCR [15-18] were selected from patients with encephalitis or respiratory complaints, including: cerebrospinal fluid (CSF, n=4), brain biopsies (n=3), nasopharyngeal swabs (n=3), nasal washings (n=1), bronchoalveolar lavage (n=1), and a plasma sample (n=1). RT-PCR panel results and Cq-values are included in the result section. The pathogens in the 13 datasets are depicted in Table 2.

For samples processed at the Great Ormond Street Hospital, London (GOSH), mRNA from the three brain biopsy samples was sequenced on an Illumina NextSeq500 instrument using an 81 bp paired-run after library preparation using Illumina’s TruSeq Stranded mRNA LT sample preparation kit (p/n RS-122-2101) according to the manufacturer’s instructions [19]. The other samples were spiked with Equine Arteritis Virus (EAV) and Phocid Herpes Virus (PhHV) internal controls preceding total nucleic acid extraction using the MagNAPure 96 DNA and Viral NA Small Volume Kit (Roche Diagnostics, Almere, the Netherlands) and sequenced on Illumina NextSeq500 (respiratory samples) or NovaSeq6000 (CSF samples, plasma) instruments using 150 bp paired-end runs after library preparation using New England BioLabs’ NEBNext Ultra Directional RNA Library preparation kit for Illumina with in-house adaptations in order to enable simultaneous detection of both DNA and RNA viruses, at the Leiden University Medical Center (LUMC) [4, 20]. Three of the CSF samples were sequenced after enrichment using capture probes targeting vertebrate viruses [21]. Human reads from the output FASTQ files were removed after mapping them to human reference genome GRCh38 [22] with Bowtie2 version 2.3.4 [23] before the datasets were uploaded to various data sharing platforms (see below).

### Data sharing

The FASTQ datasets were and remain publicly available for user-friendly downloading at https://veb.lumc.nl/CliniMG (hosted by the dept. MM, LUMC, Leiden), and part of the datasets were additionally accessible via a COMPARE Data Hub at http://www.ebi.ac.uk/ena/pathogens (hosted by the European Bioinformatics Institute, EMBL-EBI) [24].

### Bioinformatic pipelines

The datasets were analysed in a blinded fashion by the participants, with the (viral) metagenomic pipelines and classification tools (**Figure 1 and Table 1**) used at their diagnostic laboratories: Centrifuge [25], DAMIAN [26, 27], DIAMOND [28], DNASTAR [29], FEVIR [30], Genome Detective [31], Jovian [32], MetaMIC [33], MetaMix [34, 35], One Codex [36], RIEMS [37, 38], Taxonomer [39], and VirMet [40]. DAMIAN was run by two participants in combination with a different database (pipeline A and B), and one participant run both Centrifuge and GenomeDetective. Details of the algorithms are described in **Table 1**.

**Table 1.** Clinical use, classification and output characteristics of metagenomic pipelines analysed.

| Pipeline no (alphabetical order) | 1 | 2 | 3 and 3A | 4 | 5 | 6 | 7 | 8 | 9 | 10 | 11 | 12 | 13 |
| --- | --- | --- | --- | --- | --- | --- | --- | --- | --- | --- | --- | --- | --- |
| Classification tool or pipeline | Centrifuge [25] v1.0.1-beta | DAMIAN [26, 27] v190628 | DIAMOND [28] v0.9.13.114 | DNASTAR [29] Lasergene v16 | FEVIR [30] V1 | Genome Detective [31] v1.110 | Jovian [32] V0.9.6 | MetaMIC [33] v2.1.1 | MetaMix [34] [35] v1.2 | One Codex [36] v1 | RIEMS [37, 38] v4.0 | Taxonomer [39] 2020-U | VirMet [40] v1.1.1 |
| Clinical usage by participant | Patient care | Experimental | Experimental | Patient care | Patient care | Patient care<sup>a</sup> | Patient care | Patient care<sup>a</sup> | Patient care<sup>a</sup> | Experimental | Patient care | Experimental | Patient care |
| In-house/commercially available | Open-source software | Open-source software | In-house | Commercial | In-house | Commercial | In-house | In-house | In-house | Commercial | In-house | Commercial | In-house |
| Local/web-based | Local | Local | Local | Local (cloud optional) | Local | Web-based | Local | Local | Web-based (hosted on Bluebee) | Web-based | Local | Web-based | Local |
| De novo assembly | N | Y | Y [45] | N (optional) | N | Y | Y | Y | Y | N | Y | N | N |
| Alignment of NT/ AA | NT | NT, AA | AA | NT | NT | NT, AA | NT | NT | AA | NT | NT | NT, AA | NT |
| Database used by participant viral/bacterial (version) | Viruses, bacteria, fungi, archae; RefSeq (compressed index) V2019-04-04 | NCBI's nt and nr v2019-05-17, PFAM 30.0 | Viruses; NCBI's non-redundant protein database/ pipeline 3B: NCBI's nt V0.9.22 | Based on NCBI's nt V2020-01-08 | Viruses; based on Virosaurus [46] v90v_2018_11 | Viruses; based on RefSeq (filtering: Swissprot Uniref 90) v2018-11-14 | NCBI's nt, v2019-11-30 a.o. (compressed index) | Bacteria, viruses, fungi; v2.1.1 based on NCBI's nt (complete) | Human, Environmental, bacteria, Viruses RefSeq protein v2017 | Bacteria, viruses, fungi, archaea, protozoa, One Codex DB v2019-5-1 | NCBI's nt (complete) v2019-3-16 | Based on NCBI's nt (v May 2019) | Based on NCBI's nt (selection of viral full genomes without compression) v224 |
| Paired reads as input option | Y | Y | Y | Y | Y | Y | Y | Y | Y | Y | Y | N | N |
| Trimming and QC tools | Trimomatic | Trimomatic | Trimomatic [47] | Included in DNASTAR | Included in virusscan [48] 1.0 |  | Trimmomatic, fastqc, multiqc | HMN Trimmer | TrimGalore!, prinseq | Cutadapt | Roche 454 newbler | N | Seqtk, prinseq |
| Exclusion of human reads | N | Y | N/pipeline 3B:Y<sup>b</sup> | Y | N | Y | Y | Y | Y | Y | N | N | Y |
| Output | Script, | Excel | Script, | User- | Script | Web | Web | PDF | Web | Web | PDF | Web | PDF |

| type | Krona |  | krona | friendly |  | interface,<br>interactive | interface,<br>interactive |  | interface,<br>interactive,<br>PDF and<br>excel | interface,<br>interactive,<br>PDF and<br>excel |  | interface,<br>interactive |  |
| --- | --- | --- | --- | --- | --- | --- | --- | --- | --- | --- | --- | --- | --- |
| Visualizatio<br>n of<br>genome<br>coverage | N | N | N | Y | N | Y | N | N | N<br>(CLC<br>Genomics<br>Workbench) | N | Y | N (free<br>version)/ Y<br>(paid) | Y |
| Computatio<br>nal time for<br>analysis per<br>sample<br>(CPU/RAM) | ~10 min (24<br>CPUs, 0.3<br>GB RAM) | 90 min (56<br>CPUs, 125<br>GB RAM) | 90min (36<br>CPUs,<br>RAM=23 gb) | 3 min (4<br>CPUs, 64 GB<br>RAM) | 1 min (176<br>CPUs, 384<br>GB RAM) | ~10 min<br>(web-based) | 12 min (20<br>CPUs, U GB<br>RAM) | 18 min (U<br>CPUs, U GB<br>RAM) | 60-180 min<br>(web-based) | 35-40 min<br>(web-based) | U (48 CPUs,<br>768 GB<br>RAM) | 10 min<br>(web-based) | 20 min (16<br>CPUs, 64 GB<br>RAM) |
| Cut-off for<br>defining<br>positive<br>result used | ≥15 reads<br>[20] | Contig<br>length =><br>400 bp | 1 read | 1 read | ≥300 nt<br>coverage | ≥3 regions,<br>distributed<br>[49] | U | Above<br>background<br>:<br>environmen<br>tal sample | ≥3 regions<br>>10 reads<br>[50] [51]<br>Probability<br>score | 1 read | 1 read | 1 read | ≥3 reads,<br>distributed,<br>>100x than<br>NC/other<br>samples |
| Confirmato<br>ry analysis<br>required for<br>clinical<br>reporting | BLAST, PCR | PCR for<br>clinical<br>cases | BLAST, PCR<br>for clinical<br>cases | BLAST, PCR<br>for clinical<br>cases | BLAST | U | U | PCR not<br>required<br>(based on<br>validation)<br>[52-54] | BLAST,<br>coverage<br>(PCR not<br>required<br>based on<br>validation) | Not<br>required | BLAST, PCR | BLAST | U |

<sup>a</sup> Within the scope of accreditation
<sup>b</sup> Mapping of the trimmed reads to HG38 by Bowtie with “very-sensitive” option
AA; amino acid, NT; nucleotide, U; Undisclosed

**Figure 1.**
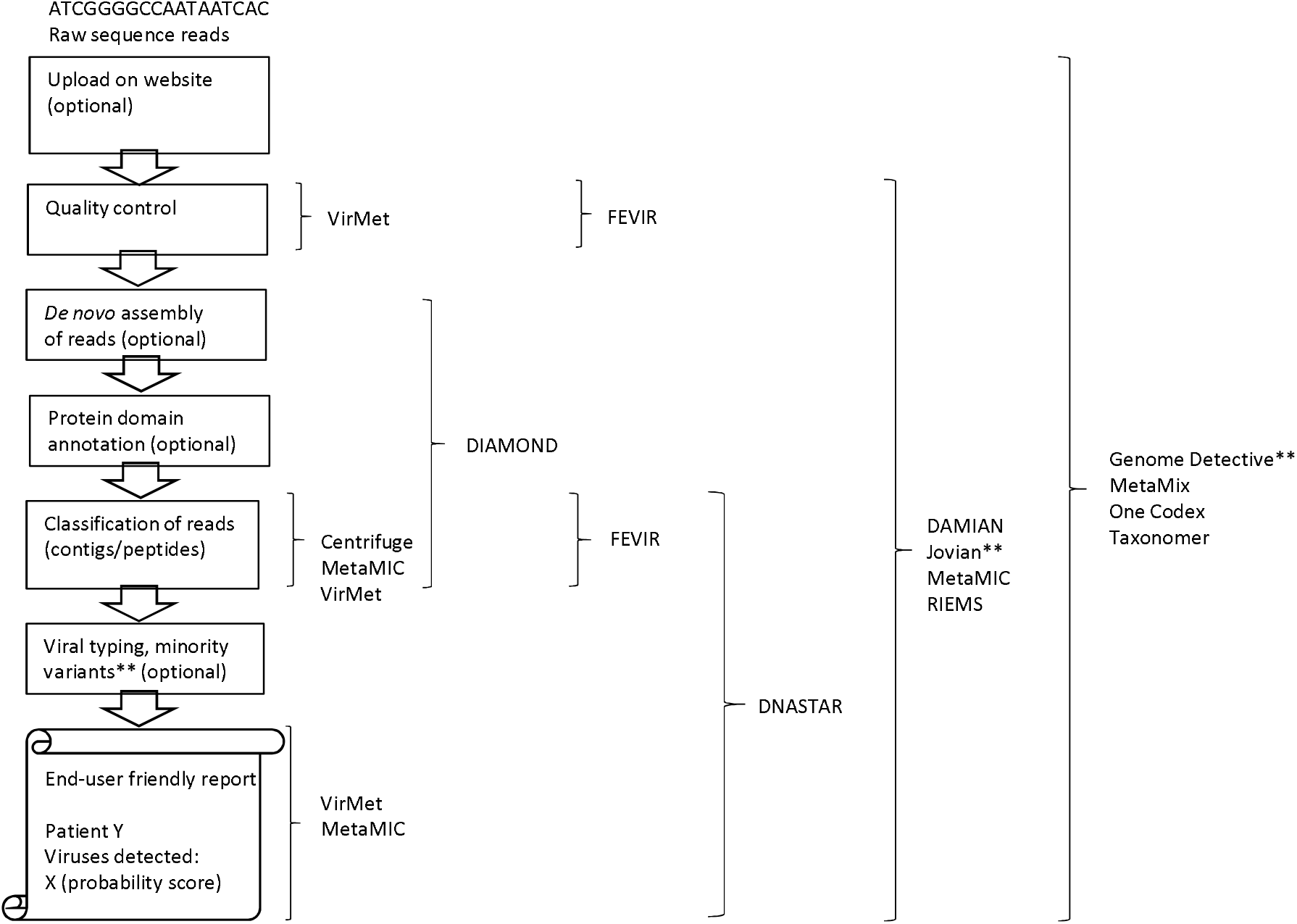
Workflow of bioinformatic analysis of (viral) metagenomics data with the pipelines and classification tools used by participants in the current study.

### Performance characteristics

Both qualitative and quantitative performance of the pipelines were analysed with real-time PCR results as gold standard. The following parameters available for all pipelines were considered: pathogen detection, taxonomic classification level and target read count. Additionally, horizontal genome coverage (if available), computational time, user-friendliness and output formats were considered. Since EAV and PhHV were added as internal controls and not reported by the participants (due to default reporting criteria, or absence in the database)they were not included in the comparative analysis.

## Results

### Metagenomic pipeline characteristics

In total 13 different metagenomic pipelines and classification tools were in use in the 13 participating diagnostic laboratories. Clinical use, classification and output characteristics of the pipelines and tools utilized are shown in **Figure 1** and **Table 1**. The majority of the pipelines were developed or adapted at a local site, while four pipelines were commercially available and web-based: DNASTAR (Madison, WI, USA), Genome Detective (Emweb bv, Herent, Belgium), One Codex (San Francisco, USA), and Taxonomer (Utah, USA). DAMIAN and Centrifuge are publicly available as an open source software. Both classification tools and reference databases differed among participants (and were fixed for end-users of the commercially available pipelines); (adapted versions of) NCBI’s nucleotide and RefSeq databases were most commonly used to generate reference databases. Six of the 13 pipelines assembled sequence reads *de novo*, whereas the others classified unassembled reads. The majority of the pipelines classified reads based on nucleotide similarity (8/13), and a minority used amino acid similarity (2/13), or a combination of both (3/13 pipelines). Parameters used by the participants for defining a positive result were the number of virus reads, horizontal genome coverage (some of the participants), and a cut-off based on posterior-probability scores of the species presence (MetaMix) and ROC-curves. Output formats varied, the majority had a user-friendly output format: excel, PDF or interactive webpage. Examples of these user-friendly output formats are shown in **Supplementary Figure S1**.

### Detection of PCR targeted viral pathogens; sensitivity

The qualitative and quantitative results of the pipeline benchmarking for viruses detected by RT-PCR are shown in **Table 2 and Figure 2**. Overall, higher abundance viral pathogens (Cq-value < 28) were detected by all metagenomic pipelines evaluated. In contrast, viral pathogens with RT-PCR Cq-value of 28 and higher including mixed virus infections were only detected by 3/13 pipelines, namely DNASTAR, FEVIR, and MetaMix. Although participants analysed the same FASTQ files, read counts of the target viruses varied from one to several orders of magnitude across pipelines. Also, read counts (all datasets combined) achieved by participants did not correlate well with the viral load as measured by RT-PCR (R=-0.07, P-value 0.5), however it must be noted that wet lab procedures varied per set of samples, including protocols with and without viral enrichment, which had potential impact on the viral read counts and thus on correlation with Cq-values. Overall sensitivity of the pipelines at sample level was 77% (10/13) - 100% (13/13 samples, mixed infections counted as one) (**Table 2 and Supplementary Table 2**). At viral mNGS hit level, overall sensitivity was 80% (12/15) - 100% (15/15 viral hits) (**Supplementary Table 4**). One of the participants reported normalized reads including the genome length, using the following formula: RPKM = (number of reads mapped to virus genome Y * 10^6^) / (total number of reads * length of genome in kp). This formula was also used to normalize the reads of all study pipelines shown in Figure 2.

**Table 2.**
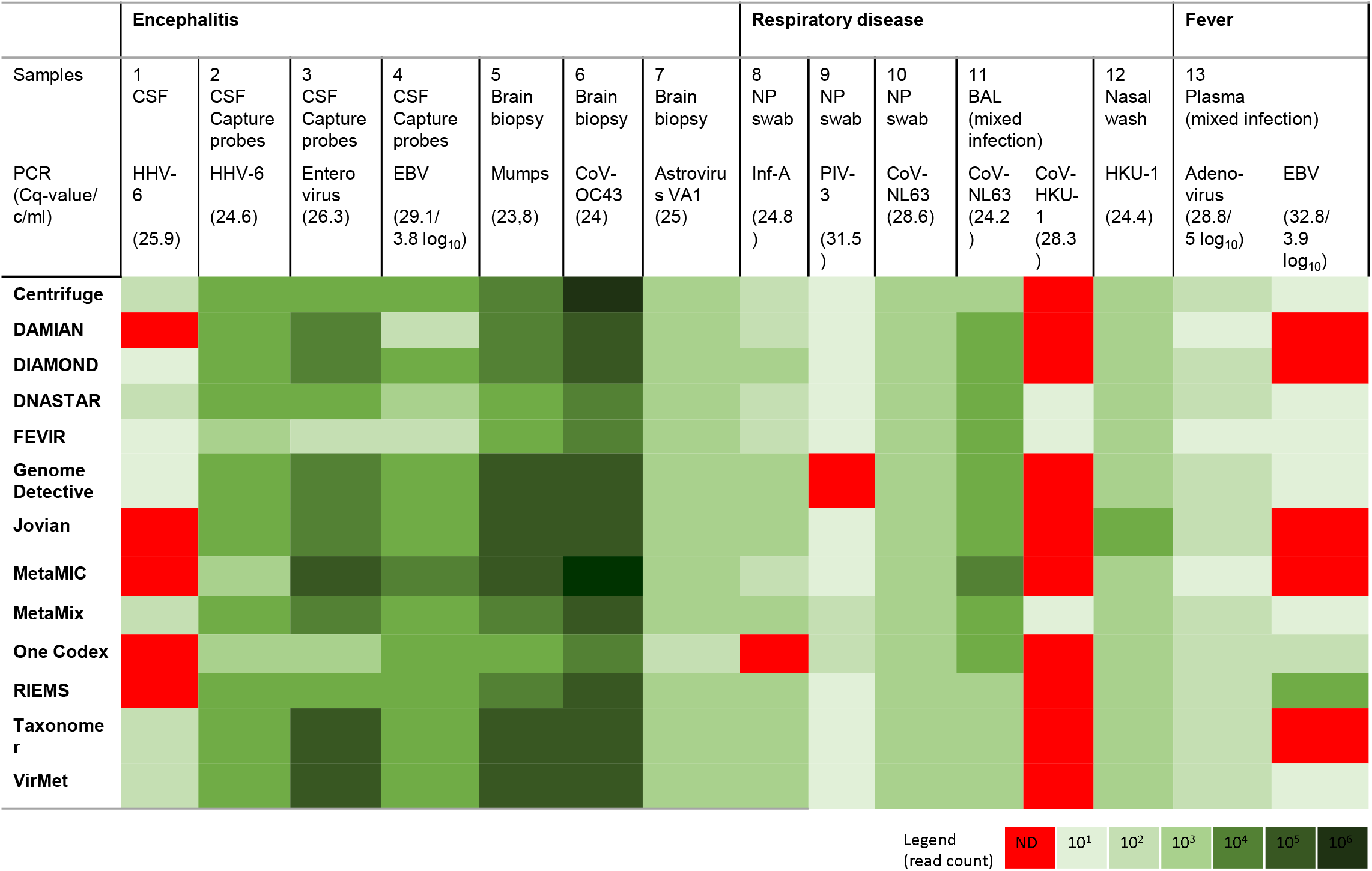
Qualitative and quantitative results: raw sequence read count categories of the PCR positive viruses reported by the metagenomic pipelines using datasets from 13 clinical samples, per classification tool (complete pipeline details can be found in table 1). CSF; cerebrospinal fluid, NP; nasopharyngeal, BAL; bronchoalveolar lavage, and in legend: ND; not detected

**Figure 2.**
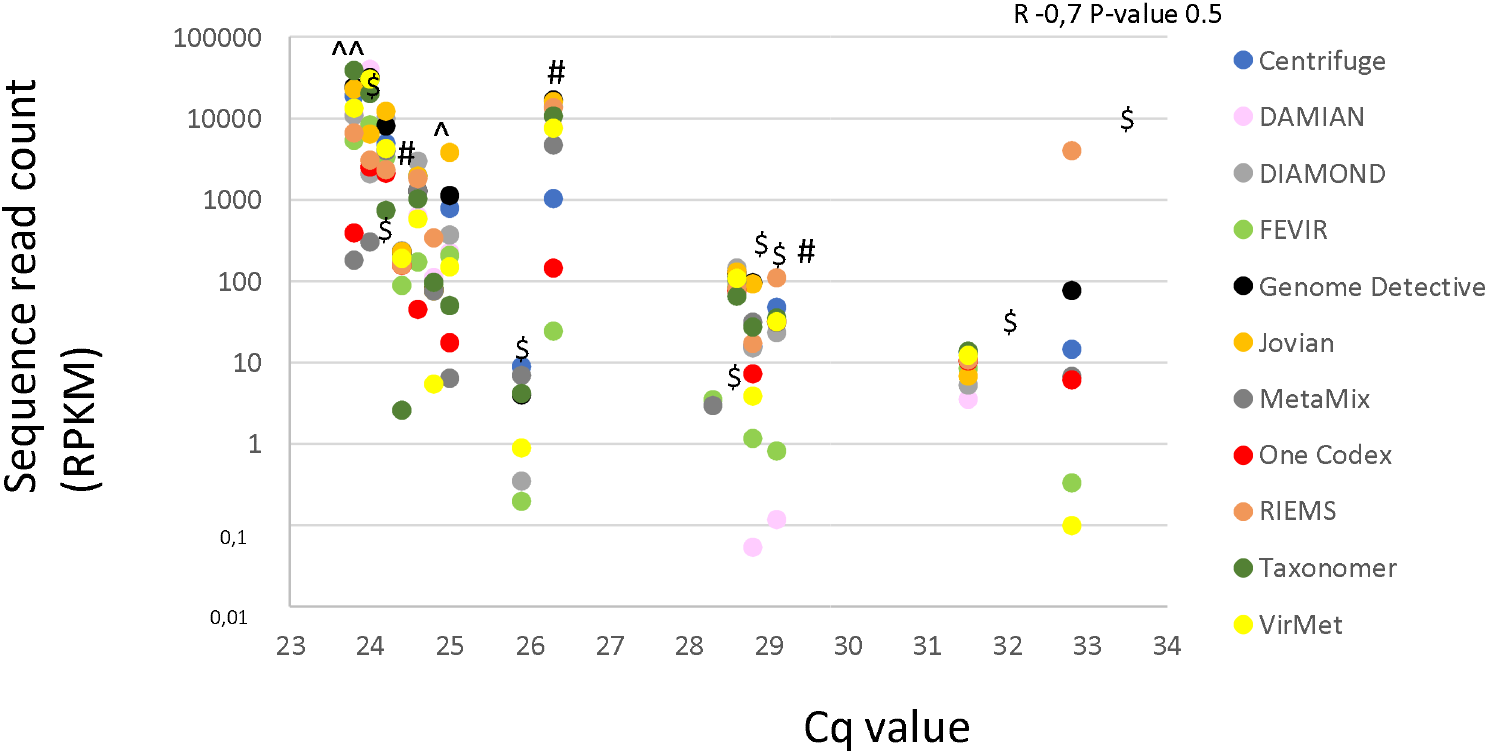
Sequence read counts (Y) versus RT-PCR Cq values (X) of the PCR positive viruses using datasets from 13 clinical samples, per classification tool with read counts reported by the participants (complete pipeline details can be found in table 1). Each vertical series of dots represents one clinical sample. The different wet lab methods used are marked (^ mRNA sequencing, $ RNA/DNA sequencing, and #: a captured approach using probes targeting vertebrate viruses). RPKM; reads per kilobase of genome per million mapped reads

### Taxonomic level of classification

The taxonomic levels of classification and typing of pathogenic viruses by the metagenomic pipelines with the settings used and reported by the participants are shown in **Figure 3** and **Supplementary Table 3**. The classification level is dependent on the database used, algorithm settings (classification of reads to the lowest common ancestor, LCA, in case of multiple hits), and the participant’s default reporting levels based on either in-house validation data or clinical relevancy. Species level classification was the most common level reported. Serotype and strain level were identified by tools that were combined with NCBI’s nt database without the LCA setting. DAMIAN was the only tool to report classification at the isolate level.

**Figure 3.**
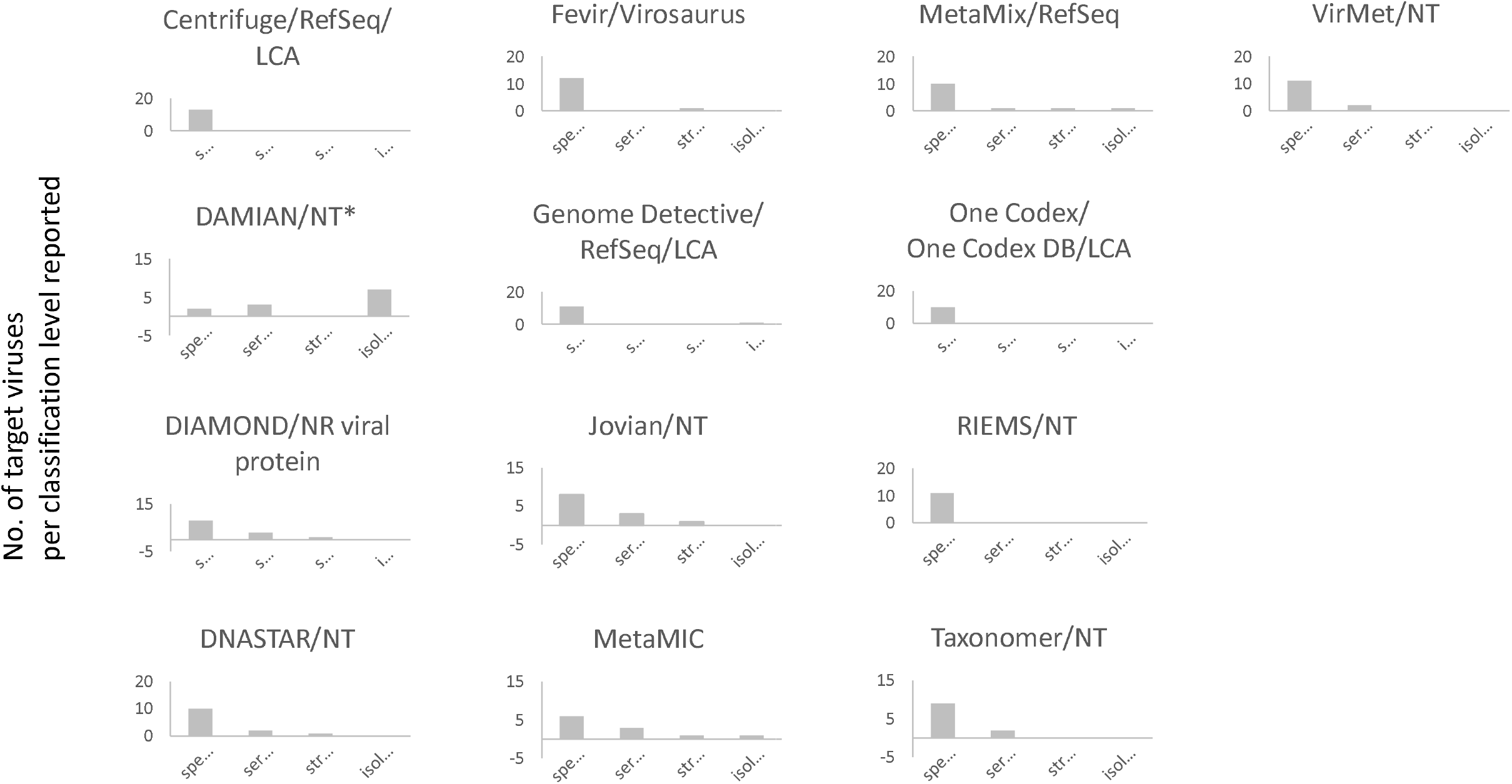
(Taxonomic) level of classification and typing of the pathogenic viruses identified using the combination of tools and databases, as reported by participating diagnostic laboratories. Depicted are the number of target viruses per classification level. RefSeq; NCBI’s RefSeq data base (or an adapted version), NT; NCBI’s nucleotide database (or an adapted version); LCA; lowest common ancestor. *Taxonomic assignment method described in [25,26]

For the Adenovirus sample (#13), virus types reported were not consistent between different pipelines: human Adenovirus type 31 (DIAMOND, Jovian, DNASTAR, VirMet), type 12 (DAMIAN), type 31 or 61 (metaMIC), indicating that type classification was not always correct. Type 12 and 31 are both from subgroup A Adenoviruses, whereas type 61 is a type 31 recombinant virus.

### Additional virus hits and positive predictive value

Additional viruses, either not tested for by RT-PCR or RT-PCR negative were reported by 11 out of 13 pipelines, and in one or more samples (**Supplementary Table 4**). The following additional viruses were reported by multiple pipelines and absent in the negative run control (dataset not available for the participants): human retrovirus RD114 (2-2102 reads, up to 28% genome coverage), feline leukemia virus (2-1406 reads), torque-teno virus (TTV) (18-66 reads, up to 7% genome coverage), polyomaviruses (5-41 reads, up to 37% genome coverage), Bovine viral diarrhea virus (BVDV) (6-220 reads, likely FBS contaminants), human metapneumovirus (HMPV) (15-21 reads, 9% genome coverage), human rhinovirus (HRV) (2-4 reads, up to 5% genome coverage), human parainfluenzavirus-4 (PIV-4) (2-6 reads) and Dengue virus (18-370 reads). RT-PCR data were available for some of the additional viruses detected (**Supplementary Table 4**). When considering viral mNGS hits with negative RT-PCR results: CoV-NL63 (1 read), PIV-4 (2-6 reads), HRV-C (2-4 reads), CoV-OC43 (5 reads), INF-B (2 reads), the positive predictive value ranged from 71-100% (**Figure 4**). It must be noted that for these mNGS hits, no distinction could be made between assignments of sequences genuinely present e.g. by index hopping (which was suspected given the low number of reads), false negative by PCR due to primers/probes mismatches, and false positive assignments. When considering the mNGS findings without available RT-PCR results, retrovirus RD114, leukemia viruses, TTV, and polyomaviruses sequences may actually be present given their association with the host (integrated or commensal).

**Figure 4.**
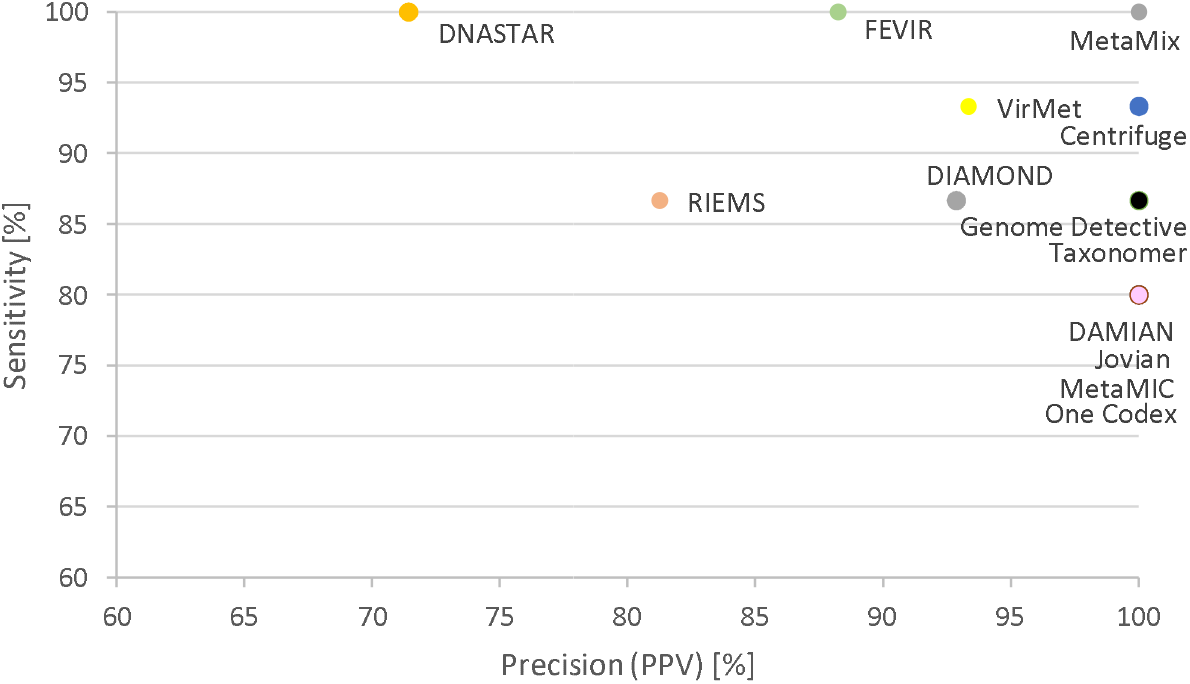
Sensitivity and positive predictive value based on the hits reported by the participants, for the different pipelines/classification tools.

### Reporting criteria

Reporting criteria used by the participants are shown in **Table 1**: a threshold for number of reads, for genome coverage (number of nucleotides and proportion of the genome, or a certain number of genome regions covered), based on reference or in-house validation studies. A BLAST analysis of matching sequences was commonly used by the study participants to exclude false positive (or to confirm true positive) hits. Some participants indicated that for clinical samples outside of the current benchmark, they required a confirmatory PCR before reporting while others indicated that this was not needed based on experiences from their validation studies.

## Discussion

This study aimed to benchmark the combination of bioinformatic tools and databases currently in use in diagnostic virology laboratories from the ESCV ENNGS network. The data presented here support bioinformatic selection and optimization of software for the implementation of viral metagenomic sequencing for pathogen detection in clinical samples. To our knowledge, this is the first large-scale international benchmarking study using datasets from clinical samples and pipelines currently applied in a large series of clinical diagnostic laboratories.

The study showed that the pipelines of all the participating laboratories succeeded in detecting viral pathogens with relative high viral loads (Cq-values <28), whereas lower abundant pathogens and mixed infections were only detected by some of the pipelines, namely DNASTAR, FEVIR, and MetaMix. These results are in line with other reports [7]. With regard to mixed infections, the less abundant viruses were generally missed, possibly due to the low number of reads, or reporting considerations. For the missed CoV-HKU1 virus, potential primer cross-reactivity with CoV-NL63 viruses was excluded by *in silico* analysis. The databases used in the pipelines were mostly custom-made, based on either NCBI’s RefSeq [41] or nt database [42]. All of the participants used different classification tools, though no selection of laboratories using different tools was made in advance. Given the inclusion of different types of pipelines including commercially available ones with fixed databases, it was not feasible to compare the different tools with one standardised database at the local sites. Two of the three pipelines that reached 100% sensitivity included NCBI’s nt database but this was also seen using a pipeline with NCBI’s RefSeq database. Pipelines with NCBI’s nt database scored both low and maximum precision. The design did allow for comparison of the complete pipeline in use for clinical diagnostics, from QC to reporting algorithms including posterior probability scores. No clear differences were observed in terms of performance based on nucleotide-based classification versus amino acid-based classification and *de novo* assembly-based algorithms versus read based classification: whereas amino-acid based classification may be more sensitive for detecting variants, two of the three pipelines with 100% sensitivity used nucleotide-based classification (DNASTAR, FEVIR). High precision was reached by pipelines that used *de novo* assembly but this was not essential: 3/8 pipelines with 100% precision did not use *de novo* assembly (Centrifuge, Taxonomer, One Codex).

Reported read counts and genome coverage varied between pipelines up to several orders of magnitude (for read counts), explaining in part the differences observed in limits of detection for samples with very low viral load. Possibly, differences in reporting of unique versus non-uniquely mapped sequence reads may be related to this difference. Sensitivity and positive predictive value were measured, conveniently avoiding the proportion of true negative findings given the immense but unknown number of negative mNGS hits without RT-PCR data needed for specificity calculations. This aspect remains a limitation intrinsically linked to mNGS validations with clinical datasets, though datasets from negative matrix samples and/or negative controls would have been contributable for specificity calculations and correction for contaminants by the participants respectively. Positive predictive value calculations were hampered by the intrinsic inability to distinguish between sequences actually present in the dataset that might be undetected by RT-PCR because, for instance, primer mismatches, index hopping or contaminant sequences introduced during library preparation.This may partially be overcome by defining mNGS consensus results as alternative golden standard, however in diagnostic settings e.g. index hopping reads should not be labelled positive despite being actually present in the dataset. A study design using synthetic datasets this may enable a more accurate estimation of the specificity and PPV *in silico* however these estimates would deviate significantly from the ones in real-life conditions, where has to be dealt with interfering factors such as the ‘kitome’, present in every single dataset. The current comparison aimed at the entire bioinformatic workflow including thresholds for reporting and corrections for interfering real-life factors.

It is important to note that participants likely have optimized their interpretation algorithm including cut-offs for their specific workflow from library preparation to sequencing. A different wet lab procedure (sequencer with or without index hopping, preparation with or without probe enrichment) will require new validation and indexing of the determined cut-off values and probability values. Because this was a dry lab comparison exercise, the participants could not follow their routine wet lab workflow and confirmatory PCR steps, which may have affected the reporting of results. Therefore no conclusions can be drawn on the limit of detection of the full metagenomic workflows used in each specific laboratory.

Genome coverage and depth was not always taken into account by the participating laboratories, however can be an effective parameter to distinguish between (PCR-)contaminants, often indicated by high depth at a small (PCR amplicon) region of the genome, and true positives [21, 55]. In five of participating laboratories a cut-off of one single read was chosen for defining a positive mNGS result. While potentially at higher risk of reporting false positive results, the PPV of these pipelines ranged from 72 up to 100%, indicating that this cut-off was dependent on the overall steps of the analysis and reporting. ROC analysis was used to find the optimal balance between sensitivity and specificity [20].

Finally, our taxonomic results are in line with data available from other groups [43]: the pipelines performed well at species level but deeper level classification was subject to less reliable classification in some cases.

In conclusion, a wide variety of viral metagenomic pipelines with overall high sensitivity are currently used in the ESCV ENNGS participating clinical diagnostic laboratories. Detection of low abundance viral pathogens or mixed infections remains a challenge, implicating the need for standardization and validation of metagenomic analysis for clinical diagnostic use [44]. The algorithm for defining positive results and rejecting false positive results is critical and should be evaluated individually for every workflow, which includes genome extraction, library preparation, sequencer and bioinformatic pipeline. Identification of deeper taxonomic levels is challenging, dependent on the individual types present in the reference database, and should be validated separately to prevent misidentification.

## Data Availability

The FASTQ datasets were and remain publicly available for user-friendly downloading at https://veb.lumc.nl/CliniMG (hosted by the dept. MM, LUMC, Leiden), and part of the datasets were additionally accessible via a COMPARE Data Hub at http://www.ebi.ac.uk/ena/pathogens (hosted by the European Bioinformatics Institute, EMBL-EBI)

## Acknowledgements

We thank the COMPARE study group (https://www.compare-europe.eu/) and the EMBL-EBI (https://www.ebi.ac.uk/) for the availability of the Data Hubs.

---

**Supplementary Figure S1.**
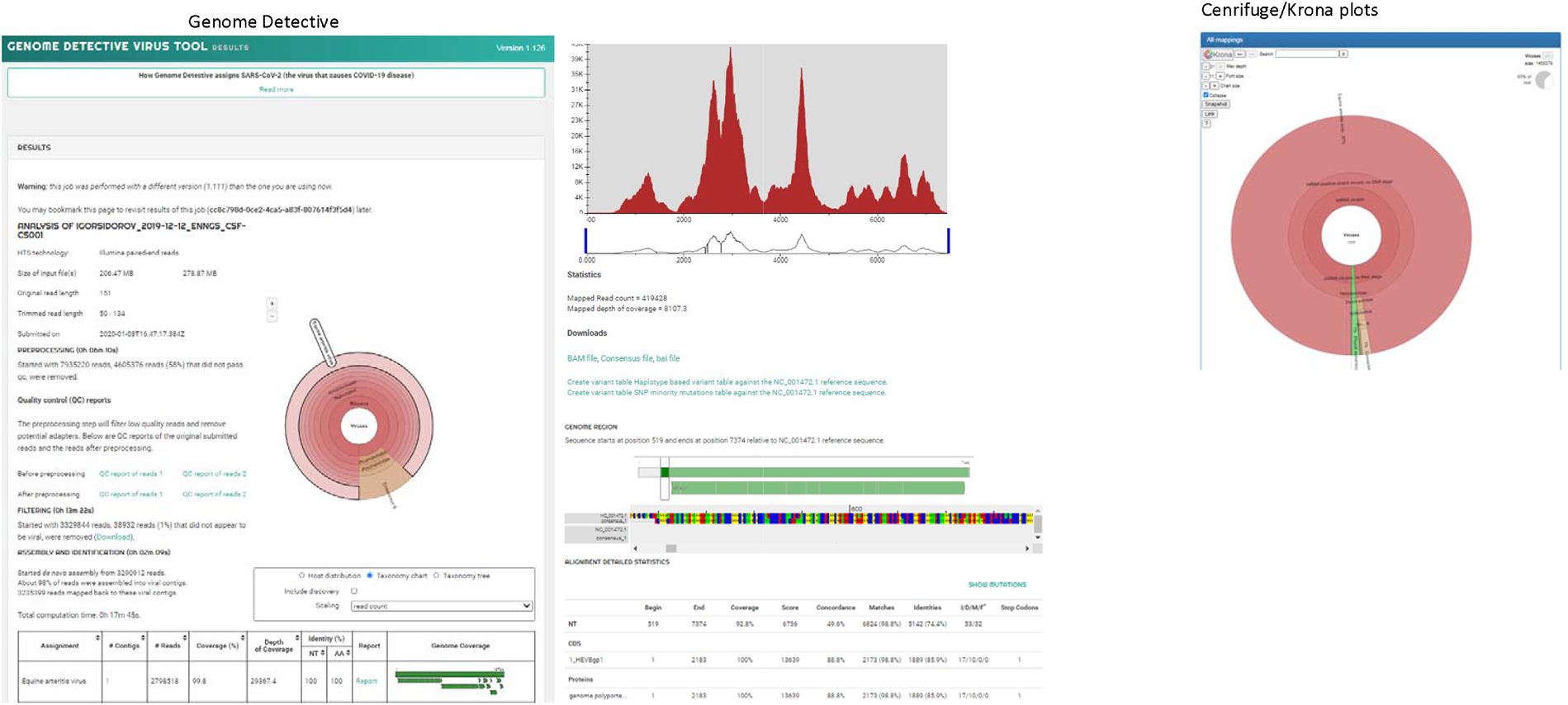
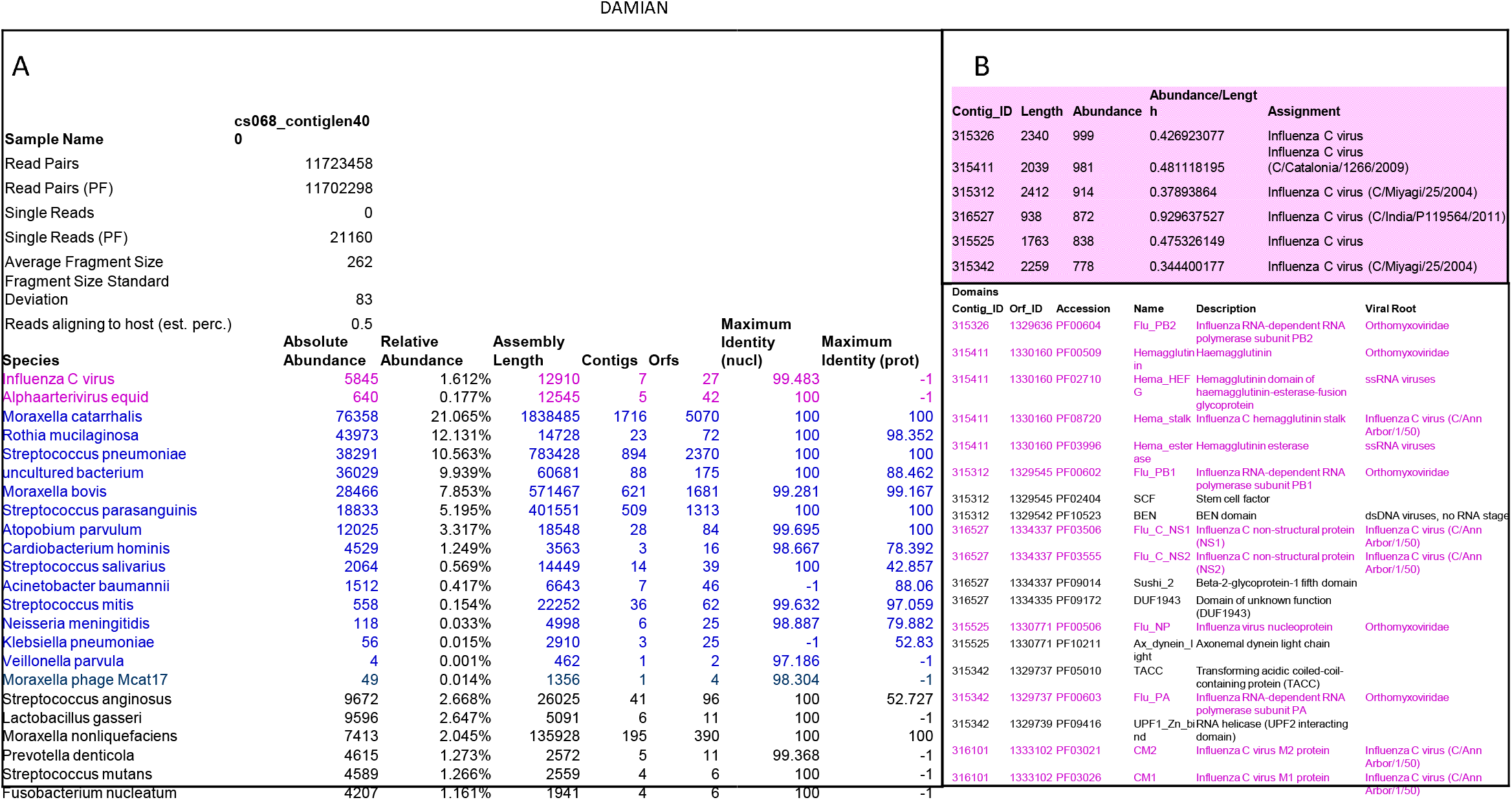
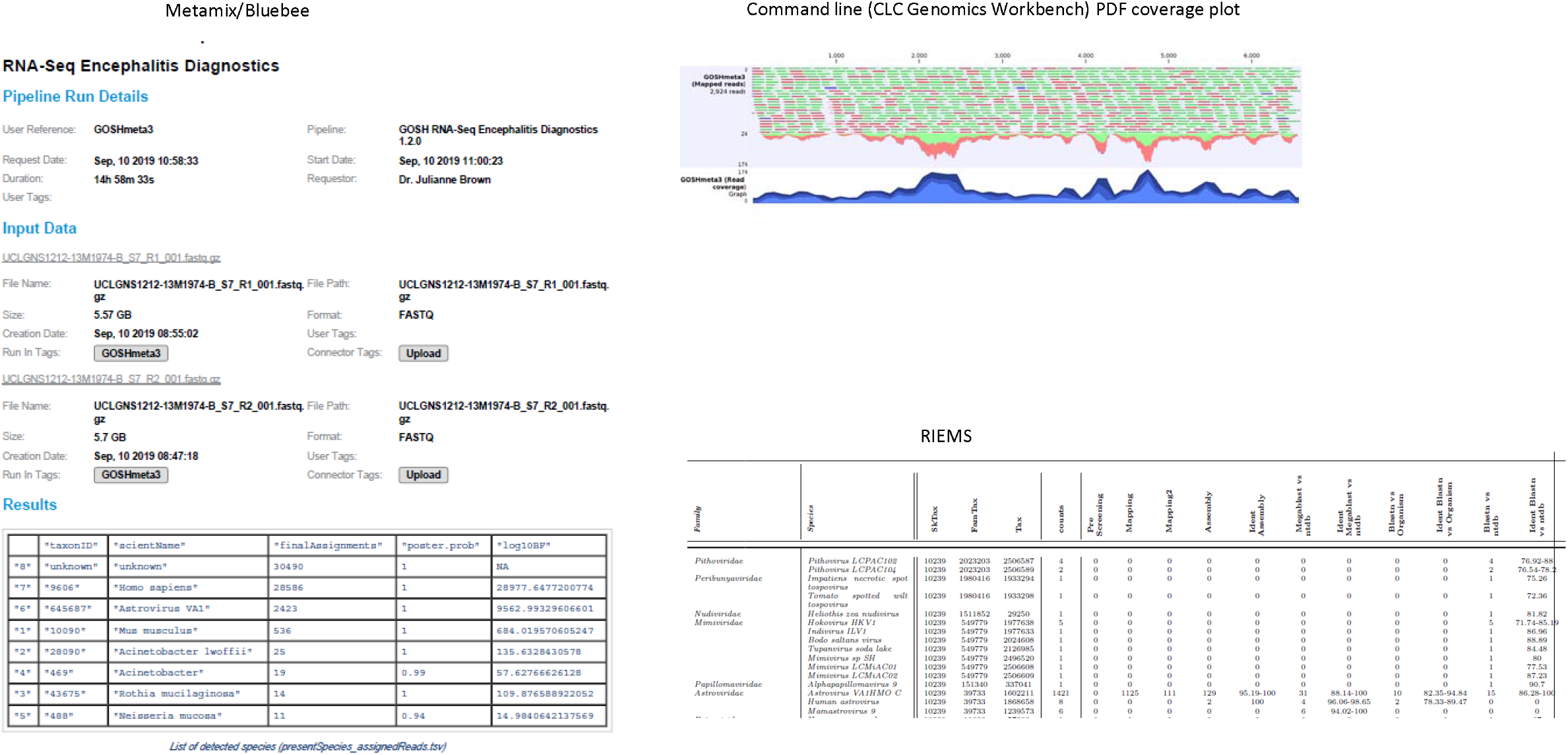
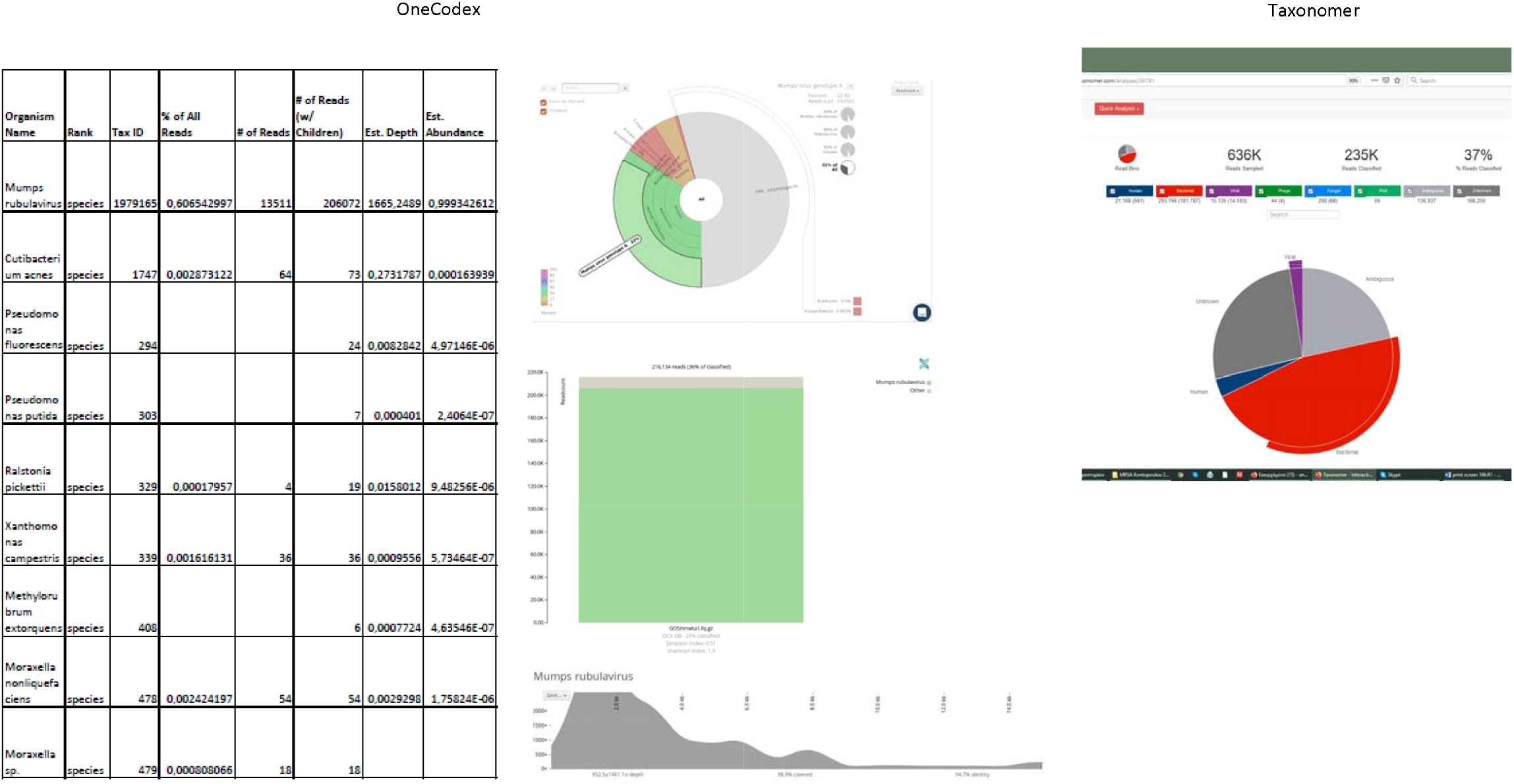
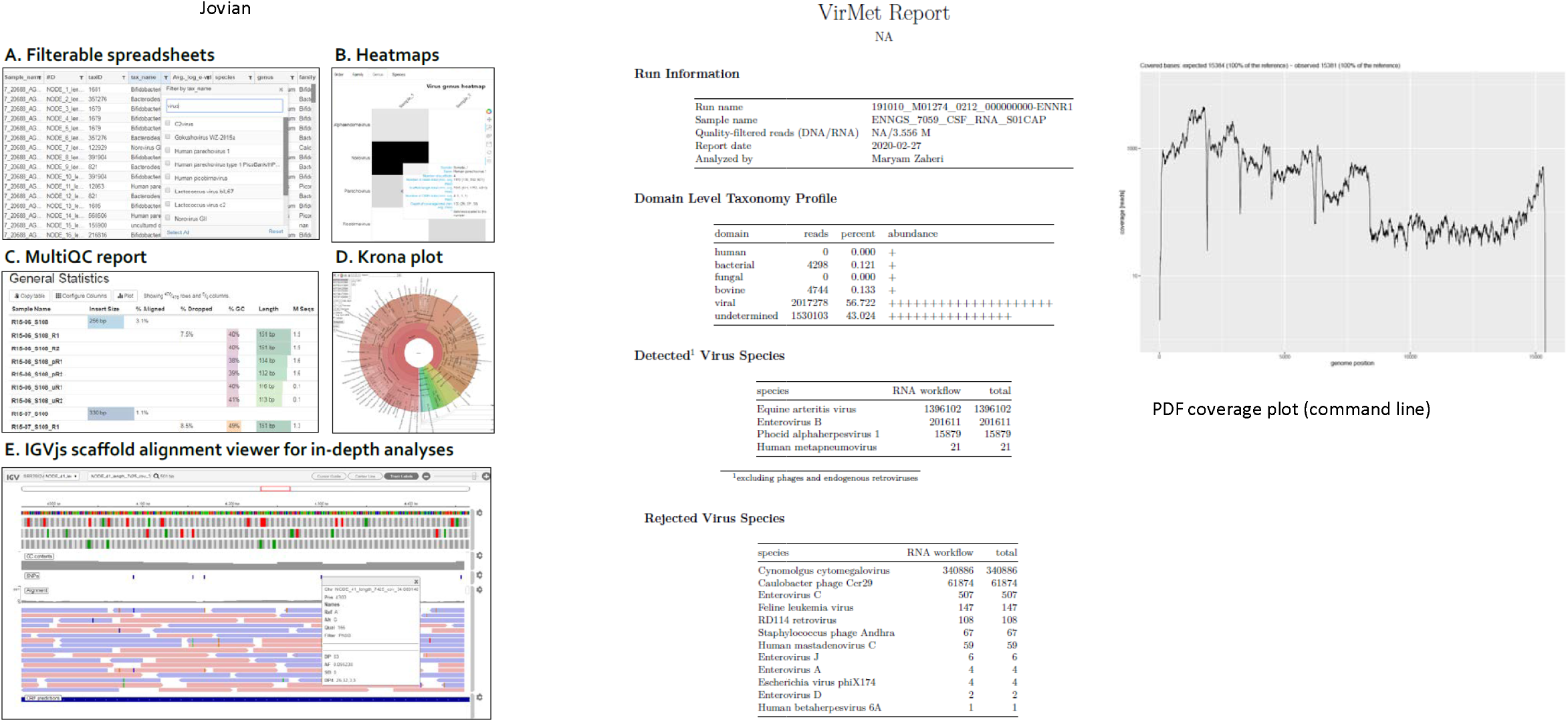
User-friendly output formats of metagenomic pipelines and tools tested, command line formats excluded.

